# Dissociable effects of dopaminergic medications on depression symptom dimensions in Parkinson’s disease

**DOI:** 10.1101/2023.06.30.23292073

**Authors:** Harry Costello, Anette-Eleonore Schrag, Robert Howard, Jonathan P. Roiser

## Abstract

**Background:** Depression in Parkinson’s disease (PD) is common, disabling and responds poorly to standard antidepressant medication. Motivational symptoms of depression, such as apathy and anhedonia, are particularly prevalent in depression in PD and predict poor response to antidepressant treatment. Loss of dopaminergic innervation of the striatum is associated with emergence of motivational symptoms in PD, and mood fluctuations correlate with dopamine availability. Accordingly, optimising dopaminergic treatment for PD can improve depressive symptoms, and dopamine agonists have shown promising effects in improving apathy. However, the differential effect of antiparkinsonian medication on symptom dimensions of depression is not known.

**Aims:** We hypothesised that there would be dissociable effects of dopaminergic medications on different depression symptom dimensions. We predicted that dopaminergic medication would specifically improve motivational symptoms, but not other symptoms, of depression. We also hypothesised that antidepressant effects of dopaminergic medications with mechanisms of action reliant on pre-synaptic dopamine neuron integrity would attenuate as pre-synaptic dopaminergic neurodegeneration progresses.

**Methods:** We analysed data from a longitudinal study of 412 newly diagnosed PD patients followed over five years in the Parkinson’s Progression Markers Initiative cohort. Medication state for individual classes of Parkinson’s medications was recorded annually. Previously validated “motivation” and “depression” dimensions were derived from the 15-item geriatric depression scale. Dopaminergic neurodegeneration was measured using repeated striatal dopamine transporter (DAT) imaging.

**Results:** Linear mixed-effects modelling was performed across all simultaneously acquired data points. Dopamine agonist use was associated with relatively fewer motivation symptoms as time progressed (interaction: β=-0.07, 95%CI [-0.13,-0.01], p=0.015) but had no effect on the depression symptom dimension (p=0.6). In contrast, monoamine oxidase-B (MAO-B) inhibitor use was associated with relatively fewer depression symptoms across all years (β=-0.41, 95%CI [-0.81,-0.01], p=0.047). No associations were observed between either depression or motivation symptoms and levodopa or amantadine use. There was a significant interaction between striatal DAT binding and MAO-B inhibitor use on motivation symptoms: MAO-B inhibitor use was associated with lower motivation symptoms in patients with higher striatal DAT binding (interaction: β=-0.24, 95%CI [-0.43,-0.05], p=0.012). No other medication effects were moderated by striatal DAT binding measures.

**Conclusions:** We identified dissociable associations between dopaminergic medications and different dimensions of depression in PD. Dopamine agonists may be effective for treatment of motivational symptoms of depression. In contrast, MAO-B inhibitors may improve both depressive and motivation symptoms, albeit the latter effect appears to be attenuated in patients with more severe striatal dopaminergic neurodegeneration, which may be a consequence of dependence on pre-synaptic dopaminergic neuron integrity.

## Introduction

Depression in Parkinson’s disease (PD) is common, affecting up to one-third of patients^1^, and is associated with greater disability^2^, increased mortality^3^, and has a greater negative impact on health-related quality of life than motor symptoms^4^. As depression in PD often occurs early in the condition and predicts increased caregiver burden^5^, greater impairment in activities of daily living^6^ and higher costs of care^7^, effective treatment of depression in PD has the potential to achieve significant health and economic benefits. However, current treatment guidelines for depression in PD advise the same approach as for depressed patients with other long-term conditions, despite evidence suggesting that standard antidepressant drugs are ineffective for these patients^8,9^.

Mood changes in PD are frequently associated with motor fluctuations, or “on/off” states, that begin as end-of-dose deterioration of the effect of dopaminergic medications, and later progress to unpredictable fluctuations^10^. One study of non-motor fluctuations in PD found that two-thirds of patients experience depressed mood exclusively during off states^11^. This raises the possibility that depression in PD may be related to dopaminergic deficit, and have a specific aetiology, explaining why treatment recommendations for depression may not generalize to PD^10^.

Depression is a heterogeneous and aetiologically complex syndrome. There are at least 256 possible unique symptom profiles that meet DSM-V criteria for a diagnosis of major depressive disorder.^12^ This degree of clinical heterogeneity has stimulated efforts to define subtypes based on symptom profiles.^12^ In PD, motivational symptoms of depression such as apathy (diminished initiation of and engagement in activities) and anhedonia (inability to experience pleasure and loss of motivation to act in order to seek pleasure) are particularly prevalent, occurring in 40% and 46% of patients, respectively^13^. Motivational symptoms are also particularly challenging to treat. The “interest-activity” symptom dimension in depression that includes loss of interest, diminished activity, fatigue, and difficulty making decisions has been associated with poor outcome of antidepressant treatment in large prospective clinical studies.^14^ People with PD report worse apathy and anhedonia when off dopaminergic medication^15,16^ and loss of dopaminergic innervation of the striatum is associated with emergent motivational symptoms in PD.^17^

Dopamine activity plays a crucial role in goal-directed behaviour, signalling how much better or worse an event is than predicted (a “reward prediction error”), as well as the valuation of reward and effort costs of actions.^18–20^ The effect of dopamine signalling depends on the dynamics of its release and clearance from the synapse.^21^ Short-latency phasic firing of dopaminergic neurons in the striatum is thought to encode reward prediction errors, crucial for reinforcement learning, while tonic levels of activity are thought to signal average reward valuation.^22^ Understanding how dopamine signalling regulates adaptive behaviour and motivation at different timescales and in different brain regions has important implications for the mechanisms underlying motivation and different depressive symptoms, and their diverse responses to different dopaminergic agents.^18,21,22^

Prescription of different dopaminergic medications in PD is currently largely driven by consideration of motor symptoms and side-effect profile, including impulse control disorders that most commonly occur with dopamine agonists. However, dopaminergic medications are also known to have antidepressant effects. Double-blind randomised controlled trials have shown that dopamine agonists can improve depression and apathy in patients with PD, and pramipexole, a relatively selective D3 receptor agonist, has shown promise as a treatment in chronic and severe depression in patients without PD.^23,24^ Lab-based studies in both animals and humans utilising dopamine depletion have consistently implicated mesolimbic dopamine activity as a key modulator of motivation.^25^ However, it remains unclear whether the antidepressant effects of dopamine agonists are primarily caused by improvement in motivational deficits rather than mood. Therefore, we aimed to perform the first comprehensive analysis of the dimensional symptom predictors of antidepressant effects of dopamine agonists.

Other treatments, such as monoamine oxidase inhibitors (MAO-Is), are also clinically used for both their antidepressant and antiparkinsonian effects.^26^ Selective type-A monoamine oxidase (MAO-A) inhibitors, such as moclobemide, are primarily used in depression to prevent the metabolism of serotonin and noradrenaline.^26^ In contrast type-B monoamine oxidase (MAO-B) inhibitors, such as rasagiline and selegiline, were developed for treatment in PD due to their ability to inhibit dopamine metabolism, thereby increasing striatal dopamine levels.^26^ However, MAO-B inhibitors such as selegiline have also been shown to improve depression in PD^27,28^ and, at high doses, they are non-selective, additionally affecting serotonergic transmission which may represent a complementary pathway to improving mood.^26,29^ There remains limited understanding of the effects of MAO-Is on different symptom dimensions of depression in PD, and no study has examined how these effects change over time or their relationship with progression of neurodegeneration within dopaminergic pathways.

We hypothesised that there would be dissociable effects of dopaminergic medications on different depressive symptom dimensions. Considering lab-based findings of the behavioural effects of dopamine agonists, we predicted that dopamine agonist treatment would improve motivational symptoms, but not other depression symptom dimensions; whereas MAO-B inhibitors may improve mood due to their concurrent serotonergic action at higher doses.^30^ Our secondary hypothesis was that dopaminergic medications with mechanisms of action reliant on pre-synaptic dopamine neuron integrity, such as MAO-B inhibitors, would have attenuated antidepressant effects as pre-synaptic dopaminergic neurodegeneration progressed.

## Materials and methods

We used data from the Parkinson’s Progression Markers Initiative (PPMI) cohort, an international multicentre cohort study.^31^ Launched in 2010, PPMI enrolled untreated, newly diagnosed PD patients and age- and sex-matched healthy controls. All participants underwent a standard battery of assessments including the MDS-Unified Parkinson’s Disease Rating Scale (MDS-UPDRS), the 15-item Geriatric Depression Scale (GDS-15) and single photon emission computed tomography (SPECT) dopamine transporter (DAT) imaging with Ioflupane [123-I] yearly, over five years.

In order to avoid capturing chronic symptoms in the context of depression with onset prior to the development of PD, all participants who had received a diagnosis of major depressive disorder more than five years prior to diagnosis with PD were excluded (n=11).

We investigated two different depression symptom dimensions within the GDS-15 previously validated in independent cohorts using factor analysis – a three-item ‘motivation’ factor^32^, and a nine-item “depression” factor^33^ (Table 1).

**Table 1.** GDS-15 depression symptom factors.

| **Table 1. GDS-15 depression symptom factors** |  |
| --- | --- |
| **“Motivation” factor** |  |
| **GDS item 2:** | Have you dropped many of your activities or interests? Scale yes (1) / no (0) |
| **GDS item 9:** | Do you prefer to stay at home, rather than going out and doing new things? Scale yes (1) / no (0) |
| **GDS item 13:** | Do you feel full of energy? Scale yes (0) / no (1) |
| **“Depression” factor** |  |
| **GDS item 1:** | Are you basically satisfied with your life? Scale yes (0) / no (1) |
| **GDS item 3:** | Do you feel that your life is empty? Scale yes (1) / no (0) |
| **GDS item 5:** | Are you in good spirits most of the time? Scale yes (0) / no (1) |
| **GDS item 7:** | Do you feel happy most of the time? Scale yes (0) / no (1) |
| **GDS item 8:** | Do you often feel helpless? Scale yes (1) / no (0) |
| **GDS item 11:** | Do you think it is wonderful to be alive? Scale yes (0) / no (1) |
| **GDS item 12:** | Do you feel pretty worthless the way you are now? Scale yes (1) / no (0) |
| **GDS item 14:** | Do you feel that your situation is hopeless? Scale yes (1) / no (0) |
| **GDS item 15:** | Do you think that most people are better off than you are? Scale yes (1) / no (0) |

A variable recording whether patients were taking a specific medication class at each timepoint was created for each of the following PD medication classes: dopamine agonists; MAO-B inhibitors; levodopa; amantadine; and catechol-O-methyltransferase (COMT) inhibitors.

Striatal DAT imaging was used as a measure of pre-synaptic dopaminergic neurodegeneration, indexing the established decrease in striatal DAT specific binding ratio (SBR) in PD as the disease progresses, owing to a loss of presynaptic dopaminergic projections from the substantia nigra and ventral tegmental area.

### Statistical analysis

We used linear mixed-effects modelling to examine the relationship between GDS-15 factor score (dependent variable) and medication state for each class of PD medication (independent variables, all included in the same multiple regression), which were acquired contemporaneously, and how this relationship changed over the progression of illness. This involved fitting both the main effects of medication class state (across all time-points) and their interactions with time. This allowed inter-individual heterogeneity and unequal follow-up intervals to be accommodated by incorporating random effects. Random intercept terms at the participant level were tested.

The interaction term between medication class state and time allowed us to assess how the relationship between each GDS-15 factor and specific medication changed over time, using all available data.

Two sets of regressions for each GDS-15 factor were conducted: 1) unadjusted and 2) adjusted for age, sex, years of education, duration of PD (all at baseline); plus cognition (Montreal Cognitive Assessment), impulsivity (Questionnaire for Impulsive-Compulsive Disorders in Parkinson’s Disease), severity of motor symptoms (MDS-UPDRS part III score, ‘off’ medication), stage of disease/functional disability (Hoehn and Yahr scale), levodopa equivalent dose (LED), antidepressant medication status and the other GDS-15 factor to ensure specificity (all at each contemporaneous timepoint). Model fit was tested using the Akaike information criterion (AIC).

Secondary analysis was conducted to assess moderation of pharmacological effects by dopaminergic neurodegeneration, replacing the time interaction with an interaction with DAT SBR. The interaction term between medication class state and DAT SBR enabled analysis of how the relationship between each GDS-15 factor and specific medication changed with striatal pre-synaptic dopaminergic neurodegeneration.

All statistical analyses were performed in R version 4.1.2. The R package ‘lme4’ was used for mixed effects modelling.

## Results

### Participant characteristics

In total, 412 participants with PD were included at baseline with loss to follow-up of one-quarter of participants by year five (Table 2). Over half of patients had commenced PD medication by the end of year one. Levodopa, dopamine agonists and MAO-B inhibitors were comparable in the prevalence of their use in year one, but, by year five, levodopa was the most common class of PD medication, with over 80% of patients prescribed this drug.

**Table 2.** Characteristics of PPMI participants with Parkinson’s disease.

|  |  |  |  |  |  |  |
| --- | --- | --- | --- | --- | --- | --- |
|  | **Baseline** | **Year 1** | **Year 2** | **Year 3** | **Year 4** | **Year 5** |
| **n** | 412 | 384 | 367 | 355 | 335 | 304 |
| **Age at entry** | 61.77 (9.76) | 61.66 (9.86) | 61.66 (9.86) | 61.64 (9.86) | 61.40 (9.96) | 60.94 (9.84) |
| **Age at diagnosis** | 61.22 (9.73) | 61.1 (9.83) | 61.1 (9.84) | 61.09 (9.82) | 60.86 (9.92) | 60.39 (9.83) |
| **Duration of PD at cohort entry (years)** | 6.56 (6.39) | 6.64 (6.51) | 6.70 (6.56) | 6.65 (6.54) | 6.57 (6.48) | 6.6 (6.53) |
| **Years of education** | 15.54 (2.99) | 15.47 (2.9) | 15.6 (2.87) | 15.61 (2.92) | 15.62 (2.90) | 15.61 (2.97) |
| **Male (%)** | 273 / 412 (66%) | 255 / 384 (66%) | 244 / 367 (66%) | 235 / 355 (66%) | 225 / 335 (67%) | 205 / 304 (67%) |
| **QUIP** | 0.29 (0.64) | 0.17 (0.47) | 0.29 (0.69) | 0.34 (0.72) | 0.39 (0.87) | 0.41 (0.81) |
| **MDS-UPDRS III** | 20.93 (8.80) | 25.25 (10.92) | 27.62 (11.39) | 29.58 (12.35) | 30.97 (12.20) | 31.27 (12.28) |
| **UPDRS total** | 32.41 (13.14) | 39.59 (16.12) | 43.02 (16.97) | 46.33 (18.80) | 48.70 (19.68) | 49.96 (19.04) |
| **MOCA** | 27.13 (2.28) | 26.28 (2.82) | 26.27 (3.14) | 26.37 (3.01) | 26.42 (3.57) | 26.53 (3.53) |
| **GDS-15** | 2.29 (2.43) | 2.53 (2.92) | 2.58 (2.87) | 2.58 (2.79) | 2.59 (2.84) | 2.78 (2.80) |
| **GDS >5** | 55 / 412 (13%) | 61 / 384 (16%) | 63 / 366 (17%) | 58 / 355 (16%) | 58 / 333 (17%) | 61 / 303 (20%) |
| **“Motivation” GDS-15 factor** | 1.02 (0.89) | 1.11 (0.97) | 1.16 (1.00) | 1.12 (1.02) | 1.19 (1.00) | 1.24 (1.04) |
| **“Depression” GDS-15 factor** | 0.74 (1.47) | 0.86 (1.80) | 0.90 (1.78) | 0.88 (1.75) | 0.86 (1.74) | 0.96 (1.73) |
| **% commenced on any PD medication** | 0/412 (0%) | 225/382 (59%) | 310/367 (84%) | 329 (355) 92.7% | 319/332 (96.1%) | 293/304 (96.4%) |
| **% commenced dopamine agonist** | - | 91/382 (23.8%) | 136/367 (37.1%) | 153/355 (43.1%) | 188/332 (56.6%) | 175/304 (57.6%) |
| **% commenced levodopa** | - | 93/382 (24.4%) | 167/367 (45.5%) | 222/355 (62.5%) | 252/332 (75.9%) | 253/304 (83.2%) |
| **% commenced MAO-B** | - | 102/382 (26.7%) | 136/367 (37.1%) | 148/355 (41.7%) | 141/332 (42.5%) | 130/304 (42.8%) |
| **% commenced amantadine** | - | 27/382 (7.1%) | 47/367 (12.8%) | 56/355 (15.8%) | 60/332 (18.1%) | 59/304 (19.4%) |
| **% commenced COMT inhibitor** | - | 1/382 (0.3%) | 2/367 (0.6%) | 10/355 (2.8%) | 14/332 (4.2%) | 17/304 (5.6%) |
| **% on antidepressant** | 29/412 (7.0%) | 29/384 (7.6%) | 33/367 (9.0%) | 30/355 (8.5%) | 32/335 (9.6%) | 26/278 (9.4%) |
| **No. DAT scans** | 408 | 360 | 341 | 10 | 280 | 7 |
| **Striatal DAT SBR** | 2.50 (0.75) | 2.21 (0.66) | 2.07 (0.68) | 1.67 (0.54) | 1.82 (0.64) | 1.30 (0.67) |
| Mean (SD); N (%); MDS-Unified Parkinson’s Disease Rating Scale (UPDRS); Montreal Cognitive Assessment (MOCA); Geriatric Depression Scale (GDS); dopamine transporter specific binding ratio (DAT SBR); Monoamine oxidase inhibitor type B (MAO-B); Catechol-O-methyltransferase (COMT) inhibitor |  |  |  |  |  |  |

Total depression symptom scores and dimension scores both increased over time. However, only 13% of patients reported a GDS-15 score >5 at baseline (suggestive of clinical depression) and less than 10% of participants with PD were taking antidepressant medication across all years of follow up, though indication for treatment was not available.

In almost all (98.8%) patients, DAT imaging was available at baseline and in years one, two and four, with only 17 participants imaged in years three and five. As reported previously in this sample, DAT SBR in PD participants was on average around half that of healthy controls; and as expected there was evidence of a marked decline over time (mean±SD percentage reduction from baseline: year 1=-9.7±17.4%, year 2=-16.6±17.7%, year 4=-26.6±18.4%).^17^

### Relationship between different depression symptom dimensions and medication class

We identified dissociable relationships between different PD dopaminergic medication classes and specific dimensions of depression (Table 3).

**Table 3.**
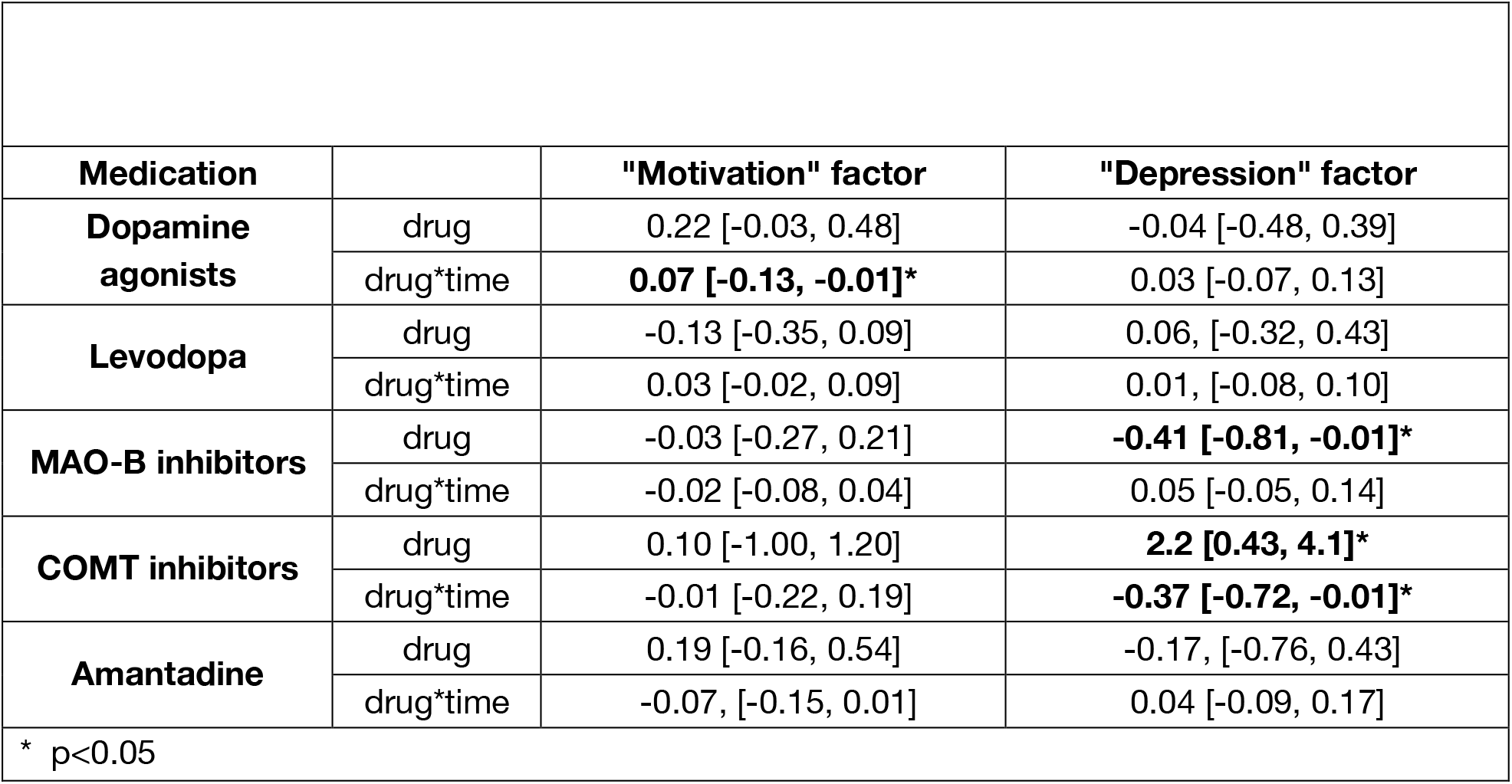
Adjusted mixed-effects model results investigating the relationship between PD medication and depression symptom dimension longitudinally (drug*year) and across all timepoints (drug)

Relationship between different depression symptom dimensions and medication class
|  |  |  |  |
| --- | --- | --- | --- |
| **Medication** |  | **"Motivation" factor** | **"Depression" factor** |
| **Dopamine agonists** | drug | 0.22 [-0.03, 0.48] | -0.04 [-0.48, 0.39] |
|  | drug*time | **0.07 [-0.13, -0.01]\*** | 0.03 [-0.07, 0.13] |
| **Levodopa** | drug | -0.13 [-0.35, 0.09] | 0.06, [-0.32, 0.43] |
|  | drug*time | 0.03 [-0.02, 0.09] | 0.01, [-0.08, 0.10] |
| **MAO-B inhibitors** | drug | -0.03 [-0.27, 0.21] | **-0.41 [-0.81, -0.01]\*** |
|  | drug*time | -0.02 [-0.08, 0.04] | 0.05 [-0.05, 0.14] |
| **COMT inhibitors** | drug | 0.10 [-1.00, 1.20] | **2.2 [0.43, 4.1]\*** |
|  | drug*time | -0.01 [-0.22, 0.19] | **-0.37 [-0.72, -0.01]\*** |
| **Amantadine** | drug | 0.19 [-0.16, 0.54] | -0.17, [-0.76, 0.43] |
|  | drug*time | -0.07, [-0.15, 0.01] | 0.04 [-0.09, 0.17] |
| * p<0.05 |  |  |  |

Longitudinal analysis revealed that dopamine agonist treatment was associated with relatively lower motivation symptoms as the disease progressed (medication-by-time interaction: β=-0.07, 95%CI [-0.13, -0.01], p=0.015), but this was not the case for depressive symptoms (medication-by-time interaction: β=0.03, 95%CI [-0.07, 0.13], p=0.6; main effect of medication: β=-0.04, 95%CI [-0.48, 0.39], p=0.8) (Figure 1). Both the model and empirical data indicated that dopamine agonist treatment was associated with lower motivation symptoms particularly in later years. However, the overall relationship between dopamine agonist treatment and motivation symptom scores across all timepoints narrowly missed significance (β=0.22, 95%CI [-0.03, 0.48], p=0.093).

**Figure 1.**
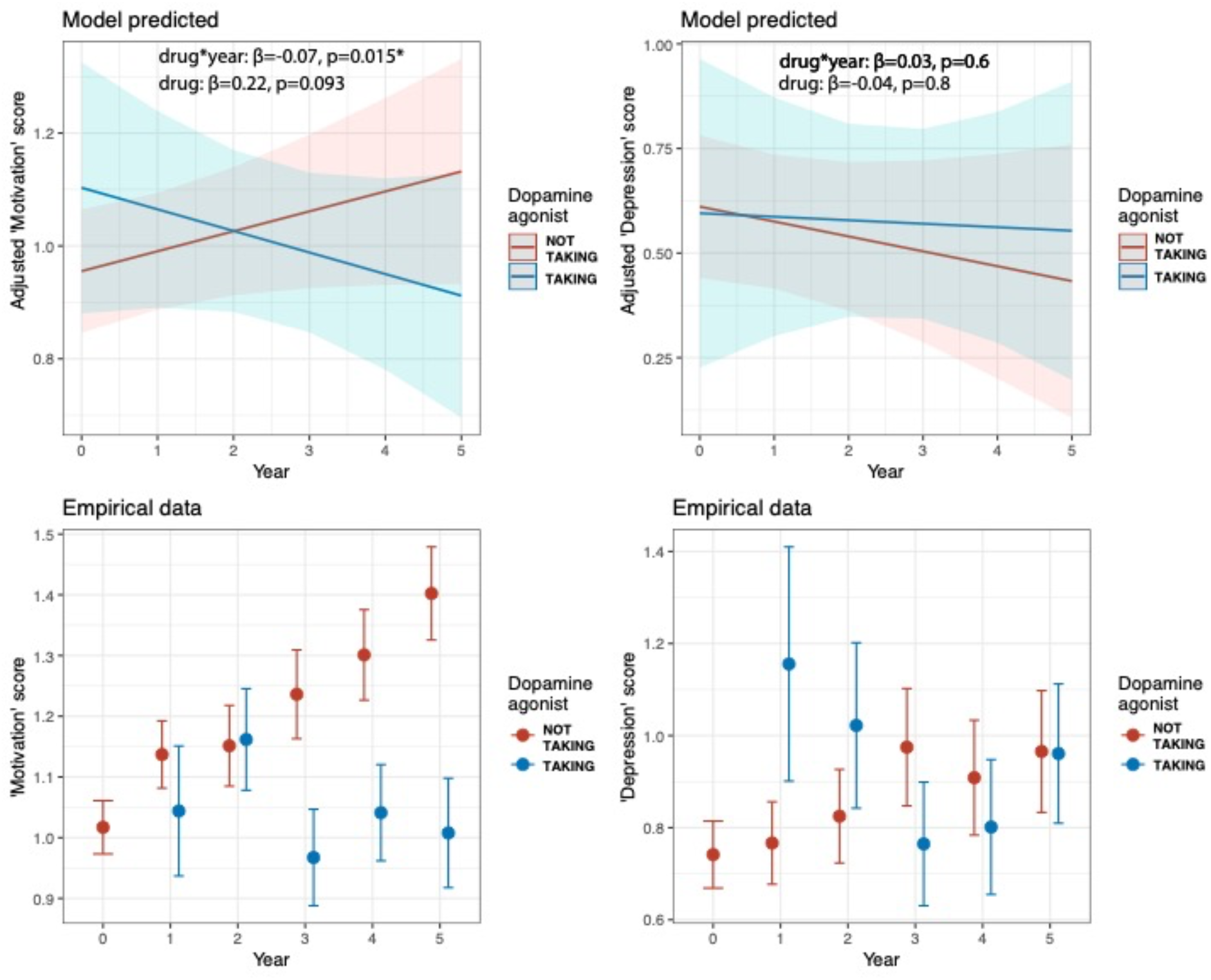
**Top row**. Adjusted mixed-effects model of the predicted relationship between dopamine agonist treatment and ‘motivation’ factor score (left)and ‘depression’ factor score (right) over time. **Bottom row**. Empirical data showing the same pattern as the modelled estimates (symptom factor score means and standard errors).

In contrast, MAO-B inhibitor treatment was associated with significantly lower depression symptoms (β=-0.41, 95%CI [-0.81, -0.01], p=0.047), but not motivation symptoms (β=-0.03, 95%CI [-0.27, 0.21], p=0.8), on average across all years of follow up. However there were no significant interactions with time (motivation symptoms β=-0.02, 95%CI [-0.08, 0.04], p=0.5; depression symptoms β=0.05, 95%CI [-0.05, 0.14], p=0.3) (Figure 2).

**Figure 2.**
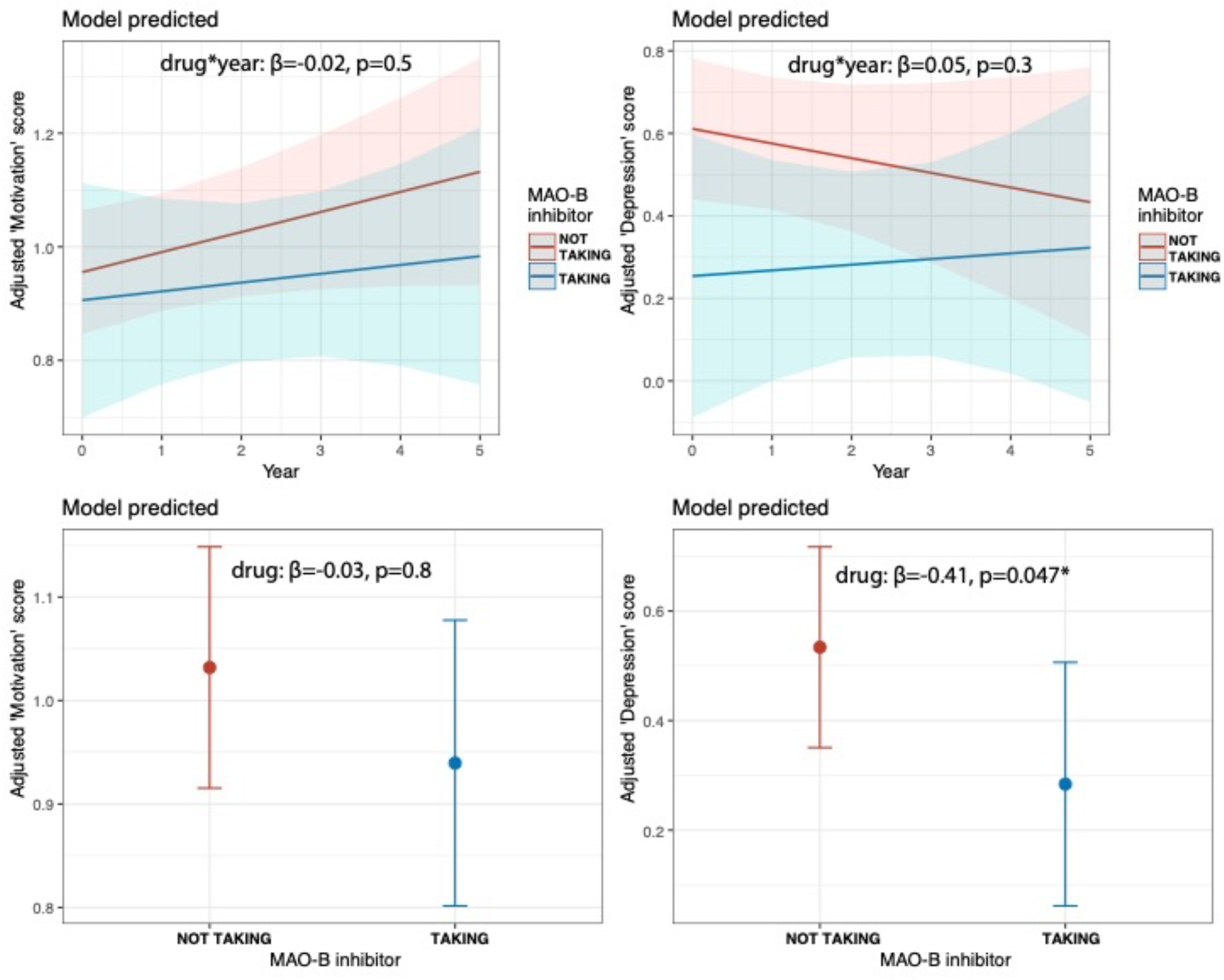
**Top row**. Adjusted mixed effects model of predicted relationship between MAO-B inhibitor treatment and ‘motivation’ factor score and ‘depression’ factor score over time. **Bottom row**. Adjusted model predicted relationship between MAO-B inhibitor treatment and ‘motivation’ factor score and ‘depression’ factor score across all timepoints.

Treatment with COMT inhibitors was associated with relatively improved depression symptoms as the disease progressed (β=-0.37, 95%CI [-0.72, -0.01], p=0.041), but worse symptoms on average across all years of follow up (β=2.2, 95%CI[0.43, 4.1], p=0.015) (Supplemental Figure S1). This inconsistent pattern of results may be a consequence of the small numbers of patients using COMT inhibitors, particularly in years 1-3 (Table 2).

No significant associations were observed between levodopa or amantadine treatment and either depression or motivation symptom factor scores (Supplemental Figure S1).

### Relationship between depression symptom dimensions, medication and striatal DAT binding

Mixed-effects model analysis of the relationship between striatal DAT SBR and depression symptoms across all timepoints revealed that the effect of MAO-B inhibitor treatment on motivation was moderated by striatal DAT SBR (β=-0.24, 95%CI [-0.43, -0.05], p=0.012) (Table 4, Figure 3). PD patients with higher striatal DAT SBR appeared to experience a greater antidepressant effect of MAO-B inhibitor treatment. A similar pattern with levodopa treatment narrowly missed statistical significance (β=-0.16, 95%CI [-0.32, 0.01], p=0.06). Conversely, if anything COMT inhibitor treatment was associated with worse motivation symptoms in patients with higher striatal DAT SBR (β=1.3, 95%CI [0.00, 2.6], p=0.05), although interpretation of this result is limited by the small number of patients taking COMT inhibitors.

**Table 4.** Adjusted mixed-effects model results investigating the relationship between PD medication and depression symptom dimension by striatal dopamine transporter (DAT) specific binding ratio (SBR)

|  |  |  |  |
| --- | --- | --- | --- |
| Medication |  | "Motivation" factor | "Depression" factor |
| Dopamine agonists | drug*DAT SBR | 0.13 (-0.09, 0.34) | 0.01 (-0.34, 0.35) |
| Levodopa | drug*DAT SBR | -0.16 (-0.32, 0.01) | -0.15 (-0.41, 0.12) |
| MAO-B inhibitors | drug*DAT SBR | **-0.24 (-0.43, -0.05)\*** | 0.02 (-0.28, 0.32) |
| **COMT inhibitors** | drug*DAT<br>SBR | 1.3 (0.00, 2.6) | -0.14 (-2.3, 2.0) |
| **Amantadine** | drug*DAT<br>SBR | 0.12 (-0.17, 0.4) | -0.02 (-0.47, 0.44) |

**Figure 3.**
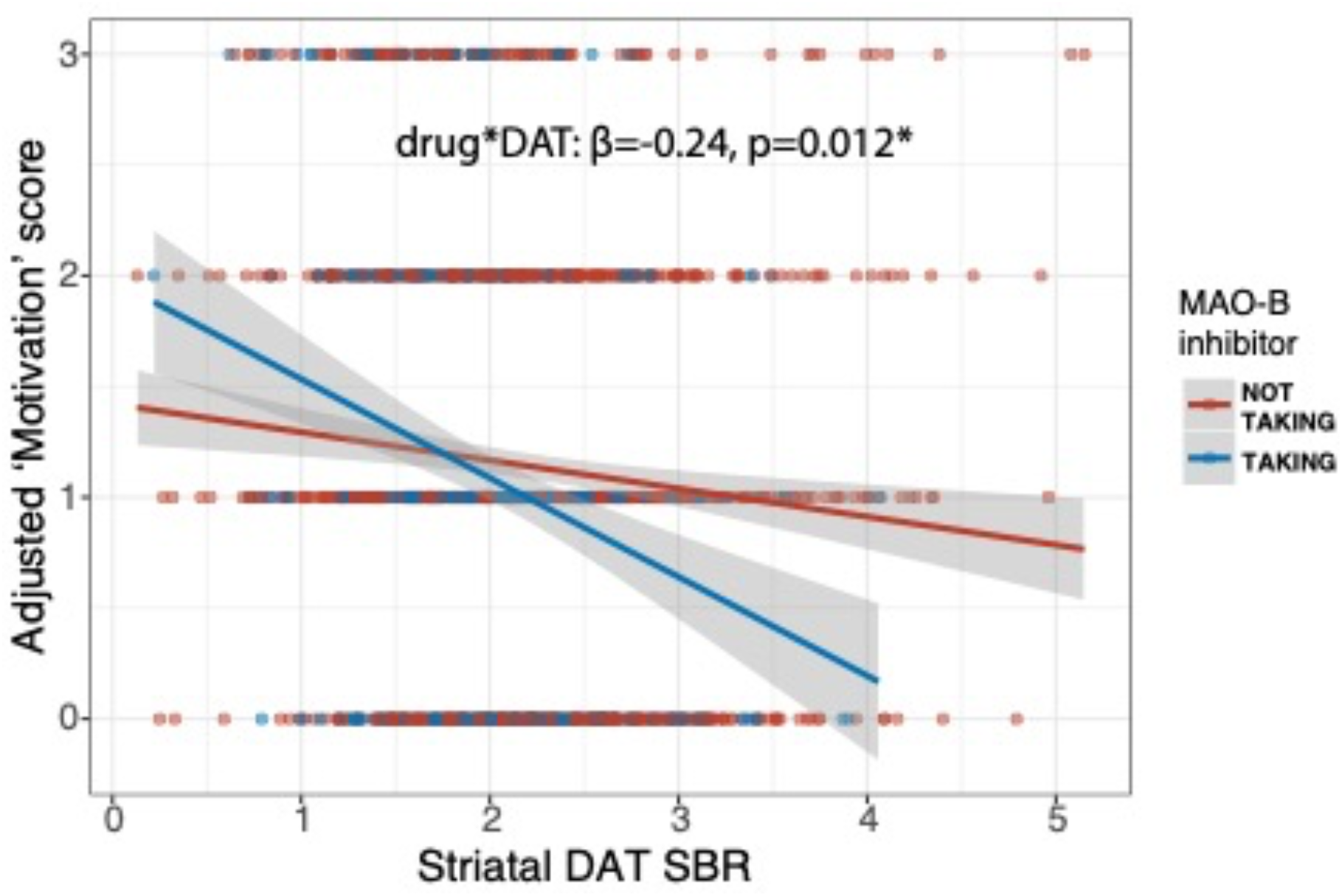
Adjusted mixed-effects model simulation showing ‘motivation’ factor score by striatal dopamine transporter (DAT) specific binding ratio (SBR) and MAO-B inhibitor treatment.

Striatal DAT SBR did not moderate the effects of dopamine agonist or amantadine treatment on individual depression symptom dimensions.

## Discussion

We performed the first longitudinal analysis of how different types of dopaminergic medication for PD are associated with different symptom dimensions of depression, and how these relationships are influenced by neurodegeneration of dopaminergic pathways. We found dissociable associations of PD dopaminergic medications with different dimensions of depression. Dopamine agonist treatment was associated with relatively lower motivation symptoms beyond the second year following diagnosis, but not depressed mood. In contrast, MAO-B inhibitor treatment was associated with fewer depression, but not motivation, symptoms. However, MAO-B inhibitor use did appear to improve motivation symptoms in patients with relatively preserved striatal pre-synaptic dopamine projections, indexed by higher striatal DAT binding. A potential explanation for this pattern is that the MAO-B inhibitor mechanism of action is dependent on pre-synaptic striatal dopamine neuron integrity, whereas other medication classes, such as dopamine agonists, act post-synaptically. Our results support existing evidence that striatal dopamine dysfunction plays a crucial role in motivation, and highlights the need for further understanding of the effect of different dopaminergic therapies on depressive symptoms as PD pathology progresses over time.^34^

Optimisation of dopaminergic therapies is often the first strategy in the treatment of depression in PD.^35^ However, at present the choice of dopaminergic therapy is largely guided by motor symptoms and potential side effects.^36^ Our findings suggest that depressive symptom profile may also have a role to play in guiding dopaminergic treatment therapy selection. For example, dopamine agonist treatment may be an effective strategy to treat motivational symptoms beyond two years following diagnosis, while alternative therapies such as MAO-B inhibitors may be indicated in patients with prominent depressed mood.

Though our study was conducted in patients with PD, these results also reveal mechanistic insights and potential treatment strategies for depression in patients without PD, specifically for those with prominent motivational symptoms that are resistant to standard antidepressant medications.^14^ The dopamine agonist pramipexole is currently used as a treatment for refractory depression and our findings support the potential for further repurposing of PD dopaminergic therapies for treatment of motivational symptoms.^37^

Dopamine agonists have been identified as an effective treatment of disorders of motivation in PD previously.^38^ However, the role of dopaminergic therapies on motivation and mood depends on dopaminergic receptor profile, pharmacodynamics, brain region and progression of underlying PD pathology.^22^ For example an animal study comparing D1-, D2- and D3-specific agonists in rescuing motivational deficits induced by lesions to the substantia nigra, found that only D3 agonists were effective^39^. D3 receptors have a narrow distribution and are primarily located within the ventral striatum, a region implicated in reward processing and the cognitive mechanisms underlying motivation. Agonism of D3 receptors has been identified as a potential treatment of apathy in PD, and further research is needed to investigate whether dopamine agonists with high affinity for D3 receptors, such as pramipexole, are more effective options for the treatment of the motivational symptoms of depression ^15,25^.

Improved understanding of the dynamics of dopamine neurotransmission underlying mood and motivation may also guide novel treatment strategies for depression in PD. Optogenetic studies have confirmed that phasic midbrain dopamine firing encodes reward prediction errors, which are crucial for learning, whereas tonic dopamine signals encode reward valuation and effort costs that drive motivation^19,40^. Dopaminergic therapies modulate the complex dynamics of dopamine signalling via inhibition of synaptic clearance mechanisms, post-synaptic agonism or potentiating synaptic release. Owing to the variation in metabolism, signalling and receptor distribution, manipulating dopamine can have paradoxical consequences for cognitive processing, often depending on basal levels of dopamine in different brain regions^22^. For example, dopamine synaptic clearance varies substantially across different brain regions. In the ventral striatum, rapid recycling via DAT predominates.^21^ In contrast, in the prefrontal cortex DAT recycling is minimal, and enzymatic degradation by catechol-O-methyltransferase (COMT) is the primary mechanism for clearance, modulating evoked dopamine release measured over minutes^18,41,42^. Though in our sample too few patients were taking COMT inhibitors to draw definitive conclusions, functional polymorphisms in COMT have been associated with motivational and mood disorders.^18,43^

Investigating the effects of different dopaminergic therapies on depression at the symptom dimension level provides insight into the potential utility of specific dopaminergic medications for the treatment of depression. However, large-scale randomised controlled trials of dopaminergic therapies for depression in PD are needed before any firm clinical recommendations can be made. Effective development of future treatments for depression in PD will depend on refining phenotypes, and integrating our understanding of the evolution of depressive symptoms over time with the progression in PD pathology.^44^

## Limitations

The associations we observed between different classes of treatment and depression symptom dimension could be a consequence of clinician treatment selection bias.

However, this is unlikely as depression symptom dimension severity did not predict treatment selection at year one.

The PPMI cohort only includes recently diagnosed patients with PD and may not be applicable to individuals in the later stages of the condition where the spread of neurodegeneration and systems involved are likely more complex.

The associations we observed between dopaminergic medication, striatal DAT binding and the depression symptom dimension could potentially be a consequence of other PD symptoms or functional disability. However, this is unlikely as all models were adjusted for cognition, motor symptoms, disease duration and functional disability.

Though all medication classes were incorporated into single model, with the sample size available in PPMI it was not feasible to account for all the interactions of the various combination therapies patients were on.

Finally, depressive symptoms were analysed as a continuous trait measure due to limited numbers of patients meeting a cut-off for clinical depression based on GDS-15 score.

Future replication of our findings in a large cohort studies of PD patients with an established clinical diagnosis of depression is needed.

## Conclusion

We identified dissociable effects of PD dopaminergic medications on different dimensions of depression. Dopamine agonists are a potentially effective treatment for motivational symptoms of depression in PD, especially beyond the second year following diagnosis. In contrast, MAO-B inhibitors may improve both depression and motivation symptoms, though the latter effect may be dependent on the severity of striatal dopaminergic neurodegeneration, potentially as a consequence of their mechanism of action depending on pre-synaptic dopaminergic neuron integrity. Further clinical trials of different dopaminergic treatment regimes are needed to identify clinically efficacious treatments for depression in PD.

## Data Availability

All data produced in the present study are available upon reasonable request to the authors

## Acknowledgements

We thank the Michael J. Fox Foundation, and the investigators, Parkinson’s patients and controls that enabled the Parkinson’s Progression Marker Initiative.

