## Supplement table S1 for "Dissociable effects of dopaminergic medications on depression symptom dimensions in Parkinson’s disease"

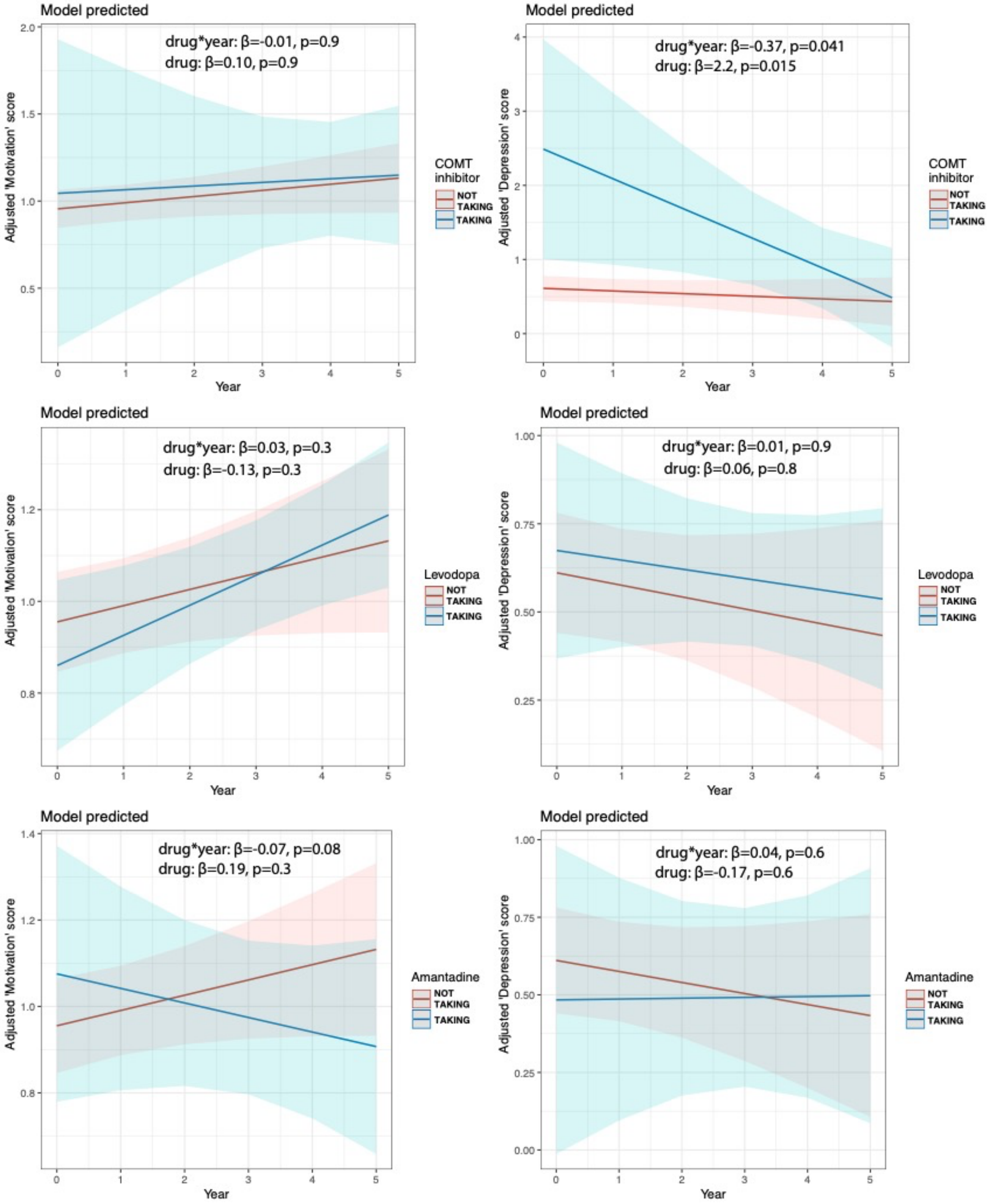


**Supplement figure S1. Top row.** Adjusted model predicted relationship between COMT inhibitor treatment and 'motivation' factor score and 'depression' factor score over time. **Middle row.** Adjusted mixed effects model of predicted relationship between levodopa treatment and 'motivation' factor score and 'depression' factor score over time. **Bottom row.** Adjusted model predicted relationship between amantadine treatment and 'motivation' factor score and 'depression' factor score over time.
